# Engagement with COVID-19 Public Health Measures in the United States: A Cross-Sectional Social Media Analysis from June to November 2020

**DOI:** 10.1101/2021.02.05.21250127

**Authors:** Daisy Massey, Chenxi Huang, Yuan Lu, Alina Cohen, Yahel Oren, Tali Moed, Pini Matzner, Shiwani Mahajan, César Caraballo, Navin Kumar, Yuchen Xue, Qinglan Ding, Rachel P. Dreyer, Brita Roy, Harlan M. Krumholz

**Affiliations:** Center for Outcomes Research and Evaluation, Yale New Haven Hospital, New Haven, Connecticut; Section of Cardiovascular Medicine, Department of Internal Medicine, Yale School of Medicine, New Haven, Connecticut; Signals Analytics, New York, New York and Netanya, Israel; Department of Sociology, Yale University, New Haven, Connecticut; Foundation for a Smoke-Free World, New York, New York; College of Health and Human Sciences, Purdue University, West Lafayette, Indiana; Department of Emergency Medicine, Yale School of Medicine, New Haven, Connecticut; Department of Medicine, Yale School of Medicine, New Haven, Connecticut; Department of Chronic Disease Epidemiology, Yale School of Public Health, New Haven, Connecticut; Department of Health Policy and Management, Yale School of Public Health, New Haven, Connecticut

**Keywords:** COVID-19 public perception, COVID-19 social media, infodemic, social media research, social media analysis, natural language processing, Reddit data, Facebook data, COVID-19 public health measures, public health surveillance

## Abstract

**Background:** The coronavirus disease 2019 (COVID-19) has continued to spread in the US and globally. Closely monitoring public engagement and perception of COVID-19 and preventive measures using social media data could provide important information for understanding the progress of current interventions and planning future programs.

**Objective:** To measure the public’s behaviors and perceptions regarding COVID-19 and its daily life effects during the recent 5 months of the pandemic.

**Methods:** Natural language processing (NLP) algorithms were used to identify COVID-19 related and unrelated topics in over 300 million online data sources from June 15 to November 15, 2020. Posts in the sample were geotagged, and sensitivity and specificity were both calculated to validate the classification of posts. The prevalence of discussion regarding these topics was measured over this time period and compared to daily case rates in the US.

**Results:** The final sample size included 9,065,733 posts, 70% of which were sourced from the US. In October and November, discussion including mentions of COVID-19 and related health behaviors did not increase as it had from June to September, despite an increase in COVID-19 daily cases in the US beginning in October. Additionally, counter to reports from March and April, discussion was more focused on daily life topics (69%), compared with COVID-19 in general (37%) and COVID-19 public health measures (20%).

**Conclusions:** There was a decline in COVID-19-related social media discussion sourced mainly from the US, even as COVID-19 cases in the US have increased to the highest rate since the beginning of the pandemic. Targeted public health messaging may be needed to ensure engagement in public health prevention measures until a vaccine is widely available to the public.

## Introduction

As the coronavirus disease 2019 (COVID-19) continues its spread in the United States (US), a key to controlling the spread until a vaccine is widely available is to enlist the public in risk-mitigation behaviors.[1,2] Studying the public’s social media posts regarding COVID-19 public health measures may provide information about targets of intervention, progress toward behavior goals, and risk of future outbreaks.[3-9] Although real-time reports on pandemic-related tests and mortality are widely available, there are fewer opportunities to gain near real-time insight into behaviors and beliefs about the pandemic.

Social media, which people are using now more than ever to communicate, has served as a useful data source in providing rapid insight into the public’s behaviors and beliefs during the pandemic.[10-13] Studies have noted a high prevalence of COVID-19-related discussion, including such topics as hygiene, shortages, and the spread of misinformation, and an increase in COVID-19-related discussion as COVID-19 cases increase.[5,14,15] However, existing findings are based on evidence during only the beginning of the outbreak, from December 2019 to April 2020, and the range of topics and keywords explored is also limited.[7,14,15-19] Additionally, studies analyzing COVID-19 behaviors and beliefs on social media have primarily used Twitter as their source, which has several limitations.[14-16] Most notably, highly rated retweets are more likely to derive from spam and bot accounts, which are also actively posting about COVID-19, and can obscure the targeting of signals from human discussions.[20-22] Further, previous studies each focused on a particular aspect of the pandemic, such as disinformation relating to the pandemic, without comparing the volume of discussion related to multiple aspects to determine the public’s relative focus on particular pandemic-related issues and behaviors. Therefore, there is a need to assess how the public’s current reaction to the pandemic has changed since the early stages, by examining broad online discussion from more diverse sources.

Accordingly, we studied changes in social media discussion of multiple topics, including daily life topics, which may be affected by the pandemic, COVID-19-related public health behavior topics, and mentions of COVID-19, from June through November 2020, and assessed their correlation with COVID-19 new daily case rates (incidence). In measuring these trends in social media data and the COVID-19 incidence rate in the US, we sought to elucidate the US public’s engagement with COVID-19-related public health measures, which are crucial to addressing the current pandemic.

## Methods

### Data Sources

The data sample consisted of unstructured, English-language posts from social media outlets and forums, such as Reddit, Facebook, and 4Chan and comments from news sites.[23] Signals Analytics, an advanced analytics consultant that conducted the analysis, accessed these data sources through a third-party data vendor, NetBase.[24,25] These social media posts were geotagged by NetBase both directly, by using geolocation data from posts, and indirectly, by using author profiles and unique domain codes (such as .uk). All data were deidentified by NetBase before being transferred to Signals Analytics.

In addition to the social data, the study included US COVID-19 case data from the COVID-19 Dashboard by the Center for Systems Science and Engineering at Johns Hopkins University.[26] These data were updated daily using a public application programming interface (API), and included total number of deaths, new daily deaths, total active cases, and daily new cases.[27]

No personal identifying information (eg, usernames, emails, or IP addresses) was shared as part of the analysis or reporting process. This study was exempted from Institutional Review Board review by Yale University as it did not engage in research involving human subjects.

### Approach

To determine trends in social media discussion during the COVID-19 pandemic, we collected data posts from all Internet sources and applied natural language processing (NLP) algorithms to identify and classify mentions of COVID-19, COVID-19-related public health measures, and daily life topics.

NetBase runs a daily query that we designed based on our project scope on over 300 million online data sources from June 15 to November 15, 2020 (eMethods 1). There were several steps to narrow the sample retrieved from the query to include only posts relevant to our research question (eTable 1). First, NLP algorithms were run to remove advertisements and pornography-related sites and posts (eMethods 2). Next, a taxonomy of topics was applied (eMethods 3). The posts that did not include discussion of topics from the taxonomy were deleted. Finally, all news articles and blog posts were deleted from the sample, so that the only remaining data posts were from social outlets (forums and comments on news sites).

**Table 1.** Number of posts by taxonomy topic (June 15 to November 15, 2020)

| Relevant Taxonomy Categories<br>(Percent Classified within All<br>Posts)<br>N=9,065,733 | Relevant Taxonomy<br>Topics | Number of Posts with Mentions<br>(Percent Classified within<br>Category) |
| --- | --- | --- |
| **COVID-19-Related Public**<br>**Health Topics**<br>**(20)** |  | **1,836,200** |
|  | Wearing Face-Mask | 1,120,344 (61) |
|  | Lockdown | 457,705 (25) |
|  | Social Distancing | 242,105 (13) |
|  | Quarantine | 94,301 (5) |
|  | Testing | 87,712 (5) |
|  | Excessive Hand-Wash | 64,679 (4) |
|  | Contact Tracing | 31,775 (2) |
|  | Reopening | 16,681 (1) |
|  | Screening | 14,569 (1) |
|  | Wearing Gloves | 11,531 (1) |
|  | Disinfection | 11,076 (1) |
|  | Wearing Face Shield | 10,104 (1) |
| **Daily Life Taxonomy Topics**<br>**(69)** |  | **6,210,255** |
|  | Sex Life | 887,457 (14) |
|  | Food | 838,513 (14) |
|  | Financial | 710,757 (11) |
|  | Travel | 651,426 (10) |
|  | Smoking/Vaping | 476,468 (8) |
|  | Mass Gatherings | 451,815 (7) |
|  | Virtual Communication | 414,549 (7) |
|  | Alcohol Consumption | 398,229 (6) |
|  | Religion | 285,538 (5) |
|  | New Skills/Hobbies Acquisition/ DIY | 280,155 (5) |
|  | Drug Use | 257,819 (4) |
|  | News/Media Consumption | 257,415 (4) |
|  | Reading | 246,074 (4) |
|  | Physical Activity | 205,116 (3) |
|  | Work from Home | 198,057 (3) |
|  | Socializing in Person | 177,522 (3) |
|  | Stockpiling | 171,421 (3) |
|  | Relaxation Techniques | 164,262 (3) |
|  | Excess Sleep | 127,623 (2) |
|  | Pets | 109,510 (2) |
|  | Postponing Plans | 98,626 (2) |
|  | Childcare | 97,735 (2) |
|  | Public Transportation | 94,414 (2) |
|  | Reduced Sleep Quality | 88,196 (1) |
|  | Home School | 80,153 (1) |
|  | Non-COVID Hospital Visits | 77,278 (1) |
|  | Doctor Well Visit | 72,235 (1) |
|  | Funerals | 45,394 (1) |
|  | Family-Centered Time | 28,106 (0) |
|  | Outdoor Culture | 21,546 (0) |
|  | Births | 17,283 (0) |
|  | Telehealth | 11,574 (0) |
|  | Smokeless Tobacco Consumption | 2,136 (0) |
| **COVID-19 Mentions (37)** |  | **3,390,139** |

The taxonomy was comprised of two categories, COVID-19-related public health measures and daily life behaviors, each of which included multiple topics (eMethods 4). COVID-19 mentions was also an individual topic in the taxonomy, independent of either category, to measure posts that directly mentioned COVID-19 by name or synonym.

Once all posts were classified according to the topics in the taxonomy, we measured trends in these topics over time by tracking the total number of posts that included mentions of each taxonomy topic and category. Classifications of topics and categories were not mutually exclusive, so the same post was able to be classified by multiple topics across any category. Trends were visualized by taxonomy category, COVID-19 mentions, and by the most commonly mentioned taxonomy topics. These trends were visualized with the COVID-19 incidence rate in the US.

This approach allowed us to identify changes in both topics that prior research in the early stage of the outbreak had shown to be prevalent in COVID-19 discussion, and in topics from daily life and COVID-19 literature reviews that were not previously known to be found in COVID-19 discussion, but may have become so or changed significantly as COVID-19 cases or current events changed.[15,16,28-32] Additionally, our approach removed redundant posts, limiting the effect of bots and reposts. The taxonomy classification was validated by calculating specificity and sensitivity (eMethods 5). In an independent data sample of 100 posts classified by manual review, the algorithm had over 80% specificity, which is higher than comparable social media research.[29] Sensitivity was calculated as the number of correct classifications of a topic using the NLP algorithms divided by the total number of posts for the topic identified by manual screening, and we found that our taxonomy approach led to an average classification rate of 92% sensitivity. We also validated the methodology by applying it to US-specific current events and found that the approach revealed an increase in online social discussion when the given current event topic was most relevant (eFigure 1). This methodology has been shown to reveal insights into outbreak characterization and event prediction for the E-cigarette or Vaping Use-Associated Lung Injury (EVALI) outbreak.[33]

**Figure 1.**
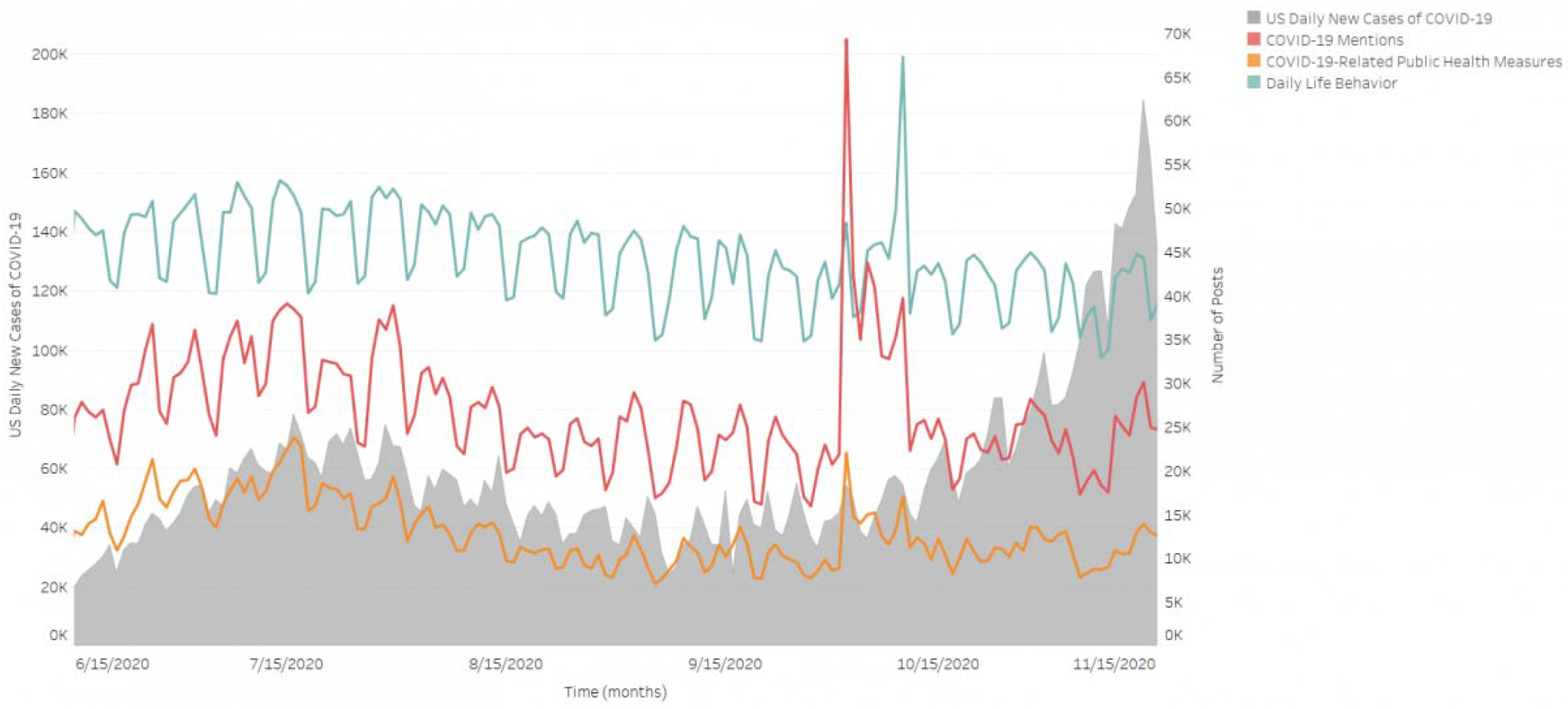
Online Social Discussion Categories vs. US Daily New COVID-19 Cases (June 15 to November 15, 2020)

## Results

The final data sample consisted of 9,065,733 online social posts that mentioned at least one of the topics in our taxonomy from June 15 to November 15, 2020 (Table 1). The majority (87%) of posts in our sample came from sources that were categorized as forums, such as Reddit and Facebook (Table 2; eTable 1).[23] The minority of posts (13%) in our sample were derived from comment sections on news sites, including The Hill, a media source focused on politics and business, and Breitbart, a right-leaning media source (Table 2; eTable 1).[34,35] Most posts in the sample were not able to be directly geotagged due to sources’ data privacy measures and restrictions. A minority were geotagged as from the US, with the remaining geotagged as from a country other than the US (eTable 2). Using indirect geotagging provided by NetBase, it was estimated that about 70% of all initial posts collected by the search query were from the US.

**Table 2.** Number of posts by source type (June 15 to November 15)

| Category of COVID-19 Discussion Topic | Posts with Mentions of COVID-19 Related Public Health Behavior | Posts with Mentions of Daily Life (%)<br>N=6,210,255 | Posts with Mentions of COVID-19 Mentions (%)<br>N=3,390,139 | Total Data Sample (%)<br>N=9,065,733 |
| --- | --- | --- | --- | --- |
|  | (%)<br>N=1,836,200 |  |  |  |
| **Forums** | 1,494,401 (81) | 5,714,446 (92) | 2,749,451 (81) | 7,928,599 (87) |
| **Comments** | 341,799 (19) | 495,809 (8) | 640,688 (19) | 1,137,134 (13) |

Within the data sample, 6,210,255 (69%) posts were classified as belonging to the category of daily life topics, 3,390,139 (37%) contained mentions of COVID-19, and 1,836,200 (20%) posts were classified as belonging to the category of COVID-19-related public health topics (Table 1). The most prevalent topics among the daily life posts were Sex Life (887,457 [14%]), Food (838,513 [14%]), and Financial Concerns (710,757 [11%]). The most prevalent topic in COVID-19-related public health behaviors posts was Wearing Face Masks (1,120,344 [61%]), followed by Lockdown (457,705 [25%]), and Social Distancing (242,105 [13%]).

Online social posts including COVID-19 mentions and COVID-19-related public health behaviors increased in June, as COVID-19 cases also increased, but remained stagnant as cases began to increase in October (Figure 1). Discussion about wearing face masks was most prevalent in mid-July, during the summer wave (mid-June to early September) of COVID-19 cases and has remained at pre-June levels in October and November, with the exception of a sharp increase on October 2, 2020 (Figure 2).

**Figure 2.**
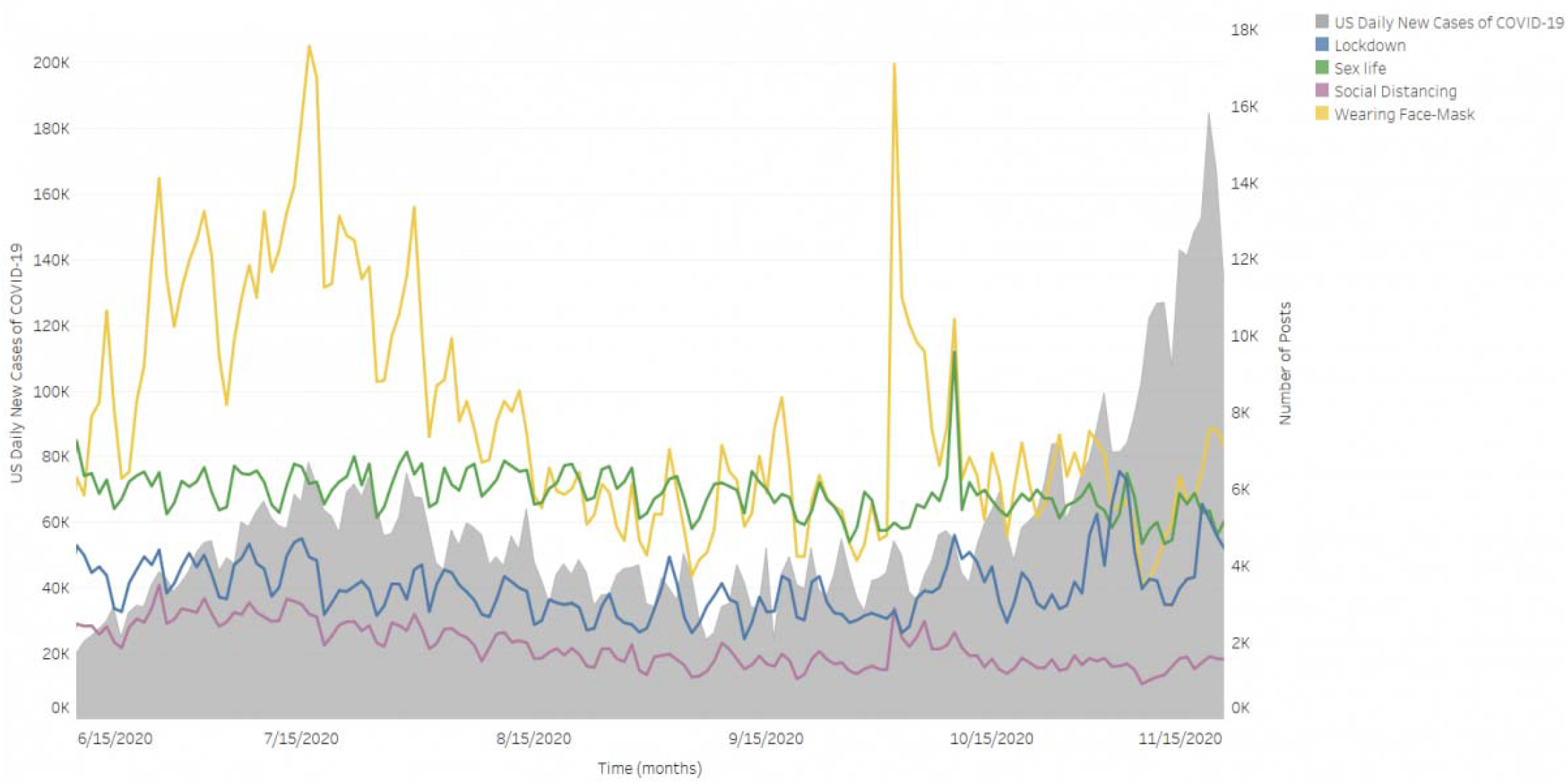
Public Health Measures Online Social Discussion vs. US Daily New COVID-19 Cases (June 15 to November 15, 2020)

## Discussion

From June to November 2020, predominantly US-based online social chatter was more focused on daily life than it was on public health behaviors relating to COVID-19. Although discussion relating to COVID-19 and related public health behaviors appeared to increase with rising US cases in the summer wave (early June to early September), the volume of COVID-19-related discussion was lower in the ongoing wave that began in the fall (mid-October), despite the fact that during the fall wave, COVID-19 cases increased to their highest rates since the pandemic began.[36] In particular, discussion of wearing facemasks, the most prevalent of any COVID-19 public health behavior we studied, declined in mid-July despite the pandemic continuing and evidence that wearing facemasks has not been universally adopted in the US, and increased only minimally once cases began to increase again in early October.[37,38] One exception was the brief but stark increase in COVID-19-related discussion on October 2, 2020, which coincided with the announcement that President Donald Trump had contracted COVID-19.[39] The high prevalence of daily life topics in social media chatter compared with COVID-19-related public health behaviors and mentions of COVID-19 is not immediately surprising given the differences in scope; however, the decrease of COVID-19 related discussion in the context of rising COVID-19 cases differs from the pattern we visualized in the summer wave and from patterns reported during the spring (March to June) wave.

Our findings differed from those of previous COVID-19-related social media analyses using Twitter and conducted earlier in the pandemic. Reports from March that used data from Twitter indicated that social media discussion about COVID-19-related health discussion was high, and more common than discussion about daily life topics such as socializing, the economy, or politics.[40] Earlier research also found that COVID-19-related public health measures were discussed more often than social topics and more often than other COVID-19-related topics.[15,7] Although this difference may be due to different sources and time periods, a change might have occurred in the public’s focus on COVID-19 preventative behaviors. For instance, as public health experts have warned against relaxing preventive behaviors as pandemic fatigue builds, activity and traffic data have indicated that people may have stopped adhering to public health recommendations to stay home and avoid close contact.[41-44] The decline of chatter regarding wearing facemasks, and the relative low rates of discussions on other COVID-19-related public health behaviors may reflect that social media engagement with these issues has decreased as the pandemic has progressed, and remains low among the US population as the pandemic continues to confront a high COVID-19 daily case rate.

Our study has several limitations. First, although our third-party data provider reported that about 70% of posts were from the US, we do not know the location for most posts according to our direct geotagging methods, which were only able to tag about 80% of posts (eTable 2). As a result, we cannot make international comparisons, but our dataset is more representative of the US than of any other country. Second, the number of posts included in our dataset was much lower than previous studies, likely due to the types of data sources used, which excluded sites such as Twitter in order to exclude noise that might have obscured signals in social media data, and our methodology, which included removing posts not relevant to our more refined taxonomy. We used a stringent exclusion criterion with a list of prespecified keywords that may also have led to a smaller sample size, but our approach aimed to create a sample with high specificity. Finally, there is no demographic information available from the data posts directly due to privacy considerations and data use agreements. Thus, we cannot determine whether our data sample contains biases due to the demographics of the people who post. For instance, Reddit, which was the most common forum source for our data sample, has been found to be used by a younger, male audience.[45,46]

## Conclusion

In this study of predominantly US-based COVID-19 social media data from June to November 2020, we observed that COVID-19 and relevant public health measures were discussed less than daily life behaviors on social media, and that discussion on wearing facemasks decreased throughout the summer and into the fall, while cases increased. These discussion rates may reveal a need for increased public health messaging as the pandemic continues.

## Data Availability

Due to the agreement between Yale and Signal Analytics, the social data used in the study are unfortunately not available to the public. However, in addition to the social data, the study included publicly-available US COVID-19 case data from the COVID-19 Dashboard by the Center for Systems Science and Engineering at Johns Hopkins University, using a public API.

https://rapidapi.com/axisbits-axisbits-default/api/covid-19-statistics/details

## Acknowledgments

Alina Cohen, Tali Moed, Pini Matzner, and Yahel Oren from Signals Analytics had full access to the data in the study and take responsibility for the integrity of the data and the accuracy of the data analysis. Daisy Massey from Yale School of Medicine takes full responsibility for the data interpretation and writing. This work was supported by the project *Insights about the COVID Pandemic Using Public Data* IRES PD: 20-005872 with funding from the Foundation for a Smoke-Free World.

## Abbreviations

COVID-19: coronavirus disease 2019
US: United States
API: application programming interface
NLP: natural language processing
EVALI: e-cigarette or vaping use-associated lung injury

## References

1. Anderson RM, Heesterbeek H, Klinkenberg D, Hollingsworth TD. How will country-based mitigation measures influence the course of the COVID-19 epidemic? The Lancet. 2020;395(10228):931–4. PMID:32164834. doi: 10.1016/S0140-6736(20)30567-5.

2. Pan A, Liu L, Wang C, Guo H, Hao X, Wang Q, et al. Association of Public Health Interventions With the Epidemiology of the COVID-19 Outbreak in Wuhan, China. JAMA. 2020;323(19):1915–23. PMID:32275295. doi: 10.1001/jama.2020.6130.

3. Signorini A, Segre AM, Polgreen PM. The use of Twitter to track levels of disease activity and public concern in the U.S. during the influenza A H1N1 pandemic. PLoS One. 2011 May 4;6(5):e19467. PMID: 21573238. doi: 10.1371/journal.pone.0019467.

4. Husnayain A, Shim E, Fuad A, Su EC-Y. Assessing the community risk perception toward COVID-19 outbreak in South Korea: evidence from Google and NAVER relative search volume. medRxiv. 2020:2020.04.23.20077552. doi: 10.1101/2020.04.23.20077552.

5. Lin Y-H, Liu C-H, Chiu Y-C. Google searches for the keywords of “wash hands” predict the speed of national spread of COVID-19 outbreak among 21 countries. Brain, Behavior, and Immunity. 2020 2020/04/10/. PMID: 32283286. doi: https://doi.org/10.1016/j.bbi.2020.04.020.

6. Puri N, Coomes EA, Haghbayan H, Gunaratne K. Social media and vaccine hesitancy: new updates for the era of COVID-19 and globalized infectious diseases. Hum Vaccin Immunother. 2020 Jul 21:1–8. PMID: 32693678. doi: 10.1080/21645515.2020.1780846.

7. Wang X, Zou C, Xie Z, Li D. Public Opinions towards COVID-19 in California and New York on Twitter. medRxiv. 2020 Jul 14. PMID: 32699856. doi: 10.1101/2020.07.12.20151936.

8. Bavel JJV, Baicker K, Boggio PS, Capraro V, Cichocka A, Cikara M, et al. Using social and behavioural science to support COVID-19 pandemic response. Nature Human Behaviour. 2020 2020/05/01;4(5):460–71. PMID: 32355299. doi: 10.1038/s41562-020-0884-z.

9. Malecki K, Keating JA, Safdar N. Crisis Communication and Public Perception of COVID-19 Risk in the Era of Social Media. Clin Infect Dis. 2020 Jun 16. PMID: 32544242. doi: 10.1093/cid/ciaa758.

10. Ella Koeze NP. The Virus Changed the Way We Internet. The New York Times. 2020 April 7, 2020. https://www.nytimes.com/interactive/2020/04/07/technology/coronavirus-internet-use.html

11. Fagherazzi G, Goetzinger C, Rashid MA, Aguayo GA, Huiart L. Digital Health Strategies to Fight COVID-19 Worldwide: Challenges, Recommendations, and a Call for Papers. J Med Internet Res. 2020 Jun 16;22(6):e19284. PMID: 32501804. doi: 10.2196/19284.

12. Li S, Feng B, Liao W, Pan W. Internet Use, Risk Awareness, and Demographic Characteristics Associated With Engagement in Preventive Behaviors and Testing: Cross-Sectional Survey on COVID-19 in the United States. J Med Internet Res. 2020 Jun 16;22(6):e19782. PMID: 32501801. doi: 10.2196/19782.

13. Brady WJ, Crockett MJ, Van Bavel JJ. The MAD Model of Moral Contagion: The Role of Motivation, Attention, and Design in the Spread of Moralized Content Online. Perspectives on Psychological Science. 2020 2020/07/01;15(4):978–1010. PMID: 32511060. doi: 10.1177/1745691620917336.

14. Lwin MO, Lu J, Sheldenkar A, Schulz PJ, Shin W, Gupta R, et al. Global Sentiments Surrounding the COVID-19 Pandemic on Twitter: Analysis of Twitter Trends. JMIR Public Health Surveill. 2020 May 22;6(2):e19447. PMID: 32412418. doi: 10.2196/19447.

15. Singh L, Bansal S, Bode L, Budak C, Chi G, Kawintiranon K, et al. A first look at COVID-19 information and misinformation sharing on Twitter. arXiv preprint arXiv:200313907. 020. PMID: 32287039

16. Abd-Alrazaq A, Alhuwail D, Househ M, Hamdi M, Shah Z. Top Concerns of Tweeters During the COVID-19 Pandemic: Infoveillance Study. J Med Internet Res. 2020 Apr 21;22(4):e19016. PMID: 32287039. doi: 10.2196/19016.

17. Massaad E, Cherfan P. Social Media Data Analytics on Telehealth During the COVID-19 Pandemic. Cureus. 2020 Apr 26;12(4):e7838. PMID: 32467813. doi: 10.7759/cureus.7838.

18. Mackey T, Li J, Purushothaman V, Nali M, Shah N, Bardier C, et al. Big Data, Natural Language Processing, and Deep Learning to Detect and Characterize Illicit COVID-19 Product Sales: An Infoveillance Study on Twitter and Instagram. JMIR Public Health Surveill. 2020 Aug 3. PMID: 32750006. doi: 10.2196/20794.

19. Mackey T, Purushothaman V, Li J, Shah N, Nali M, Bardier C, et al. Machine Learning to Detect Self-Reporting of Symptoms, Testing Access, and Recovery Associated With COVID-19 on Twitter: Retrospective Big Data Infoveillance Study. JMIR Public Health Surveill. 2020 Jun 8;6(2):e19509. PMID: 32490846. doi: 10.2196/19509.

20. Chu Z, Gianvecchio S, Wang H, Jajodia S. Who is tweeting on Twitter: human, bot, or cyborg? Proceedings of the 26th Annual Computer Security Applications Conference; Austin, Texas, USA: Association for Computing Machinery; 2010. p. 21–30. https://www.eecis.udel.edu/~hnw/paper/acsac10.pdf

21. Tsou M-H, Zhang H, Jung C-T. Identifying data noises, user biases, and system errors in geo-tagged twitter messages (Tweets). arXiv preprint arXiv:171202433. 2017.

22. Ferrara E. # COVID-19 on Twitter: Bots, Conspiracies, and Social Media Activism. arXiv preprint arXiv:200409531. 2020.

23. Bernstein M, Monroy-Hernández A, Harry D, André P, Panovich K, Vargas G. 4chan and/b: An Analysis of Anonymity and Ephemerality in a Large Online Community. 2011. https://www.aaai.org/ocs/index.php/ICWSM/ICWSM11/paper/view/2873/4398

24. Signals Analytics. [December 11, 2020]; Available from: https://www.signals-analytics.com.

25. NetBase Quid. [December 11, 2020]; Available from: https://netbasequid.com.

26. Dong E, Du H, Gardner L. An interactive web-based dashboard to track COVID-19 in real time. Lancet Infect Dis. 2020 May;20(5):533–4. PMID: 32087114. doi: 10.1016/S1473-3099(20)30120-1.

27. Axisbits. COVID-19 Statistics API Documentation. Rapid API2020. https://rapidapi.com/axisbits-axisbits-default/api/covid-19-statistics/details

28. Han X, Wang J, Zhang M, Wang X. Using Social Media to Mine and Analyze Public Opinion Related to COVID-19 in China. Int J Environ Res Public Health. 2020 Apr 17;17(8). PMID: 32316647. doi: 10.3390/ijerph17082788.

29. Jelodar H, Wang Y, Orji R, Huang H. Deep Sentiment Classification and Topic Discovery on Novel Coronavirus or COVID-19 Online Discussions: NLP Using LSTM Recurrent Neural Network Approach. IEEE J Biomed Health Inform. 2020 Jun 9;Pp. PMID: 32750931. doi: 10.1109/jbhi.2020.3001216.

30. Amelia Nirenberg AP. Schools Reopening: The State of Play for K-12. The New York Times. 2020 August 17, 2020. https://www.nytimes.com/2020/08/17/us/k-12-schools-reopening.html

31. Abby Goodnough KS. CDC Weighs Advising Everyone to Wear a Mask. The New York Times. 2020 March 31, 2020. https://www.nytimes.com/2020/03/31/health/cdc-masks-coronavirus.html

32. Sheikh K. You’re Getting Used to Masks. Will You Wear a Face Shield? The New York Times. 2020 May 24, 2020. https://www.nytimes.com/2020/05/24/health/coronavirus-face-shields.html

33. Matzner P. Using Advanced Analytics for the Early Detection of Pandemics and Outbreaks. Early Release; Signals Analytics. April 13, 2020. https://www.signals-analytics.com/resources/white-papers/early-detection-pandemics-outbreaks

34. The Hill AllSides Media Bias Rating. [October 15, 2020]; Available from: https://www.allsides.com/news-source/hill-media-bias.

35. Ribeiro FN, Henrique L, Benevenuto F, Chakraborty A, Kulshrestha J, Babaei M, et al., editors. Media bias monitor: Quantifying biases of social media news outlets at large-scale. Twelfth International AAAI Conference on Web and Social Media; 2018. https://www.aaai.org/ocs/index.php/ICWSM/ICWSM18/paper/view/17878/17020

36. Wan WD, Jacqueline. U.S. hits highest daily number of coronavirus cases since pandemic began. The Washington Post. 2020 October 23, 2020. https://www.washingtonpost.com/health/2020/10/23/covid-us-spike-cases/

37. Katz JS-K, Margot; Quealy, Kevin. A Detailed Map of Who Is Wearing Masks in the U.S. The New York Times. 2020 July 17, 2020. https://www.nytimes.com/interactive/2020/07/17/upshot/coronavirus-face-mask-map.html

38. Brenan M. Americans’ Face Mask Usage Varies Greatly by Demographics. Gallup News2020. https://news.gallup.com/poll/315590/americans-face-mask-usage-varies-greatly-demographics.aspx

39. Baker PH, Maggie. Trump Tests Positive for the Coronavirus. The New York Times. 2020 October 2, 2020. https://www.nytimes.com/2020/10/02/us/politics/trump-covid.html

40. Molla R. How coronavirus took over social media. Vox. 2020 March 12, 2020. https://www.vox.com/recode/2020/3/12/21175570/coronavirus-covid-19-social-media-twitter-facebook-google

41. Miller SW, Jane. Fauci Says U.S. Won’t Get Back to Normal Until Late 2021. NBC News. 2020 September 11, 2020. https://www.nbcnews.com/health/health-news/fauci-says-us-won-t-get-back-normal-until-late-n1239882

42. Ghader S, Zhao J, Lee M, Zhou W, Zhao G, Zhang L. Observed mobility behavior data reveal social distancing inertia. arXiv preprint arXiv:200414748. 2020.

43. Traffic Monitoring Count Data: Volume and Classification Information. 2020 [updated September 28, 2020October 7, 2020]; Available from: https://portal.ct.gov/DOT/PP_SysInfo/Traffic-Monitoring.

44. Schuman R. INRIX U.S. National Traffic Volume Synopsis Issue #15 (June 20-June 26, 2020). 2020 [updated June 29, 2020October 7, 2020]; Available from: https://inrix.com/blog/2020/06/covid19-us-traffic-volume-synopsis-15/.

45. Vasilev E. Inferring gender of Reddit users. 2018. https://kola.opus.hbz-nrw.de/opus45-kola/frontdoor/deliver/index/docId/1619/file/Master_thesis_Vasilev.pdf

46. Finlay SC. Age and gender in Reddit commenting and success. 2014. doi: 10.1633/JISTaP.2014.2.3.2

